# Extensive testing may reduce COVID-19 mortality: a lesson from northern Italy

**DOI:** 10.1101/2020.04.24.20078709

**Authors:** Mauro Di Bari, Daniela Balzi, Giulia Carreras, Graziano Onder

## Abstract

We examined data on the progression of COVID-19 epidemics in four regions in northern Italy. Lombardy, Emilia-Romagna, and Piedmont had an extremely steeper increase in mortality with increasing number of tests performed than Veneto, which applied a policy of broader swab testing. This suggests that the strategy adopted in Veneto, similar to that in South Korea, is effective in containing COVID-19 epidemics and should be applied in other regions of Italy and countries in Europe.

On February 20, 2020, a first autochthonous case of COVID-19 respiratory disease was observed in Lombardy, Italy (1), soon followed by a second patient in Veneto, which borders Lombardy. Since then, the outbreak has rapidly expanded, mostly in regions in northern Italy (2).

Initially, epidemiological surveillance and strategies for swab testing were under control of regional healthcare authorities. On February 25, the Italian Ministry of Health issued more stringent testing policies for application of swabs to identify COVID-19 cases, prioritizing patients with respiratory symptoms and possible COVID-19 contacts who required hospitalization. Most regions promptly complied with these recommendations, whereas Veneto maintained its policy, implemented after the occurrence of the first cases, of extensive testing and isolation of positive cases (3). Surprisingly, the debate stemming from these different regional policies valued international more than Italian evidences (4). We aimed at assessing, using data from the first month of the Italian experience, how different policies for swab testing may impact on the initial progression of COVID-19 epidemics.

Data were obtained from the reports of the Italian Department of Civil Protection, issued since February 24, which include daily number of swabs performed and deaths from COVID-19 in each region (5). We compared Lombardy, Emilia-Romagna, and Piedmont, three regions in northern Italy that closely followed the recommendations for restrictive COVID-19 testing, and Veneto, which applied a policy of broader testing (3).

**Conflict of interest:** None declared.

**Funding statement:** No funding.

## METHODS

The cumulative number of tests performed in the first month, from February 24 through March 27, and COVID-19 cumulative mortality from March 2 through April 3, were indexed by population in each region (6). The 7-day lag time between COVID-19 testing and mortality was allowed because death occurs usually 7+ days after clinical onset and diagnosis (2).

Piecewise linear regression was applied, separately for the four regions, to identify the breakpoints in the slope of the number of tests performed over time, and to examine whether the relationship between the cumulative number of tests performed through each date (independent) and mortality (dependent variable) followed a different progression over time in the four regions.

The effectiveness of the two testing strategies was estimated by regressing the number of tests (dependent) and the cumulative lagged mortality (independent variable), separately for the two most distant scenarios of Lombardy and Veneto: the slope of these regressions represents the number of tests associated with one death.

The proportion of positive cases was calculated as the percent ratio of the 3-day moving average of positive cases over the 3-day moving average of tests performed, to compensate for delays and imprecisions in the daily reporting of data. One-way ANOVA was used to evaluate differences in the proportion of positive cases across the four regions, applying the Games Howell test for unbalanced variances for post-hoc comparisons. Pearson’s r correlation coefficient was used to assess in each region whether the proportion of positive tests changed with the cumulative number of tests performed. Statistical significance was set at p<0.05.

## FINDINGS

**Figure 1** shows that the number of tests increased daily from February 24 through March 27 in all regions, although with marked differences. Starting from 0.3, 1.5, 0.3, and 4.5 tests per 10,000 persons by February 24, Emilia-Romagna, Lombardy, Piedmont and Veneto reached 107.2, 95.3, 45.2 and 170.5 tests per 10,000 persons by March 27. Piecewise regression showed that the progressive increase in the number of tests performed changed slope in all regions, although with different timing and extension. The slope increased by March 10 in Lombardy, March 14 in Veneto and Piedmont, and March 17 in Emilia-Romagna. It was initially lowest in Piedmont, intermediate in Lombardy and Emilia-Romagna, and maximal in Veneto (0.4, 1.3, 1.2, and 2.3 more tests per 10,000 per day, respectively). Then, it increased to a similar extent in Veneto and Emilia-Romagna, although it remained smaller in Lombardy and Piedmont than in the other two regions (8.7, 7.4, 4.2, and 2.8 more tests per 10,000 per day, respectively).

**Figure 1.**
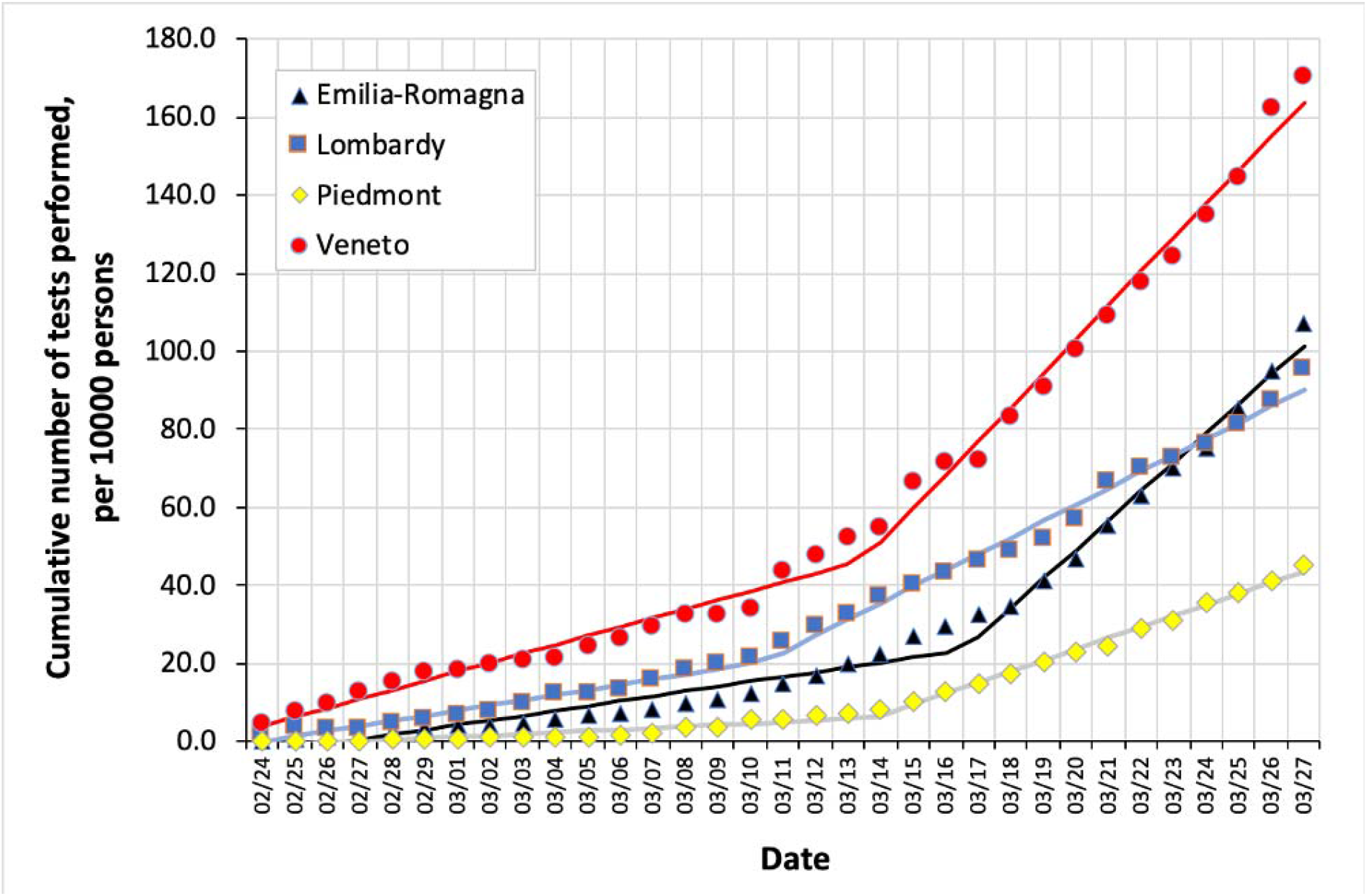
Cumulative number of COVID-19 tests performed in four regions in norther Italy from February 24 through March 27, per 10,000 persons in each region. Piecewise regression lines are also shown.

From March 2 through April 3, mortality increased from 0.02, 0.04, 0.00, and 0.004 to 4.27, 8.26, 2.39, and 1.17 per 10,000 persons in Emilia-Romagna, Lombardy, Piedmont and Veneto, respectively. **Figure 2** shows the relationship between the cumulative number of COVID-19 tests performed from February 24 through March 27, and the corresponding lagged mortality (March 2-April 3) in the four regions. Compared to Veneto, Lombardy, Piedmont and until March 17 also Emilia-Romagna clustered towards a steeper mortality rate increase with increasing number of tests performed. After that date, in Emilia-Romagna the slope of the relationship flattened substantially: the piecewise regression confirmed a change in the slope when the cumulative number of 41.1 tests per 10,000 was reached, in coincidence with the sudden increase in the rate of daily testing on March 18. Slopes remained unchanged in the other regions. When the cumulative number of tests performed was regressed linearly towards lagged mortality in Lombardy and Veneto, the slope of the regression was 133 in Veneto and 10.4 tests per one death in Lombardy.

**Figure 2.**
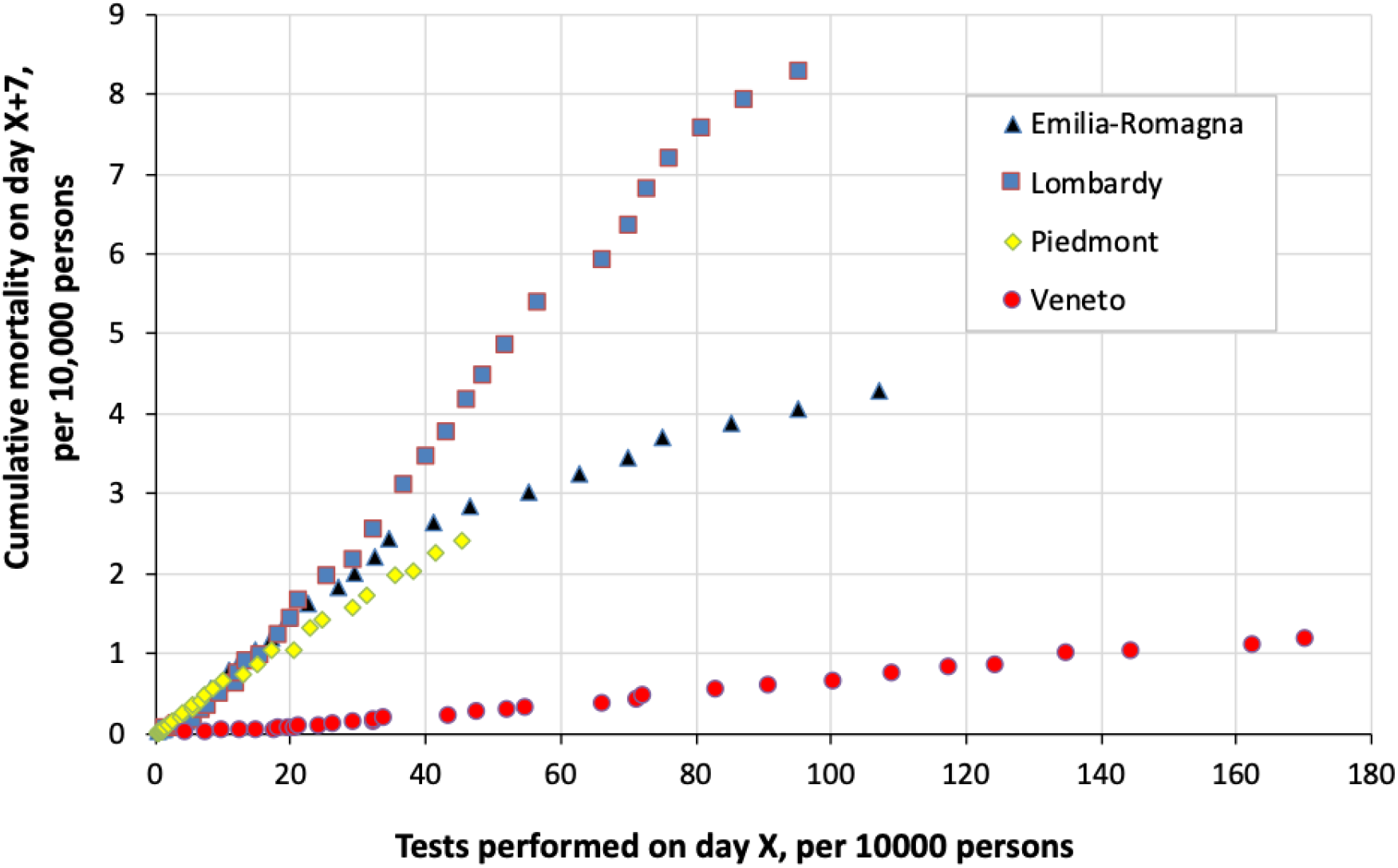
Cumulative COVID-19 mortality in four regions in norther Italy from March 2 through April 3, as a function of the cumulative number of COVID tests performed seven days before, i.e. from February 24 through March 27.

The proportion of positive tests (ratio between 3-day averaged numbers of positive tests and of tests performed) was (mean±SD) 26.8±9.4, 33.7±13.1, 28.7±12.3 and 8.0±3.2 percent in Emilia-Romagna, Lombardy, Piedmont, and Veneto, respectively (p<0.001); post-hoc paired comparisons showed significant differences (p<0.001) between Veneto and each of the other regions. The proportion of positive tests decreased with the number of tests performed in Emilia-Romagna (r=-0.381, p=0.017), remained unchanged in Lombardy (r=0.164, p=0.318) and Veneto (r=0.184, p=0.262) and increased in Piedmont (r=0.341, p=0.034).

## CONCLUSIONS

We observed that extensive swab testing, applied since the beginning of the epidemics, may reduce the spread of COVID-19, by identification of a high number of positive cases that can be eventually isolated (3,7). Four regions in the same area of Italy, with similar demographics, economics, and healthcare system, which were almost simultaneously hit by the virus, adopted different strategies for COVID-19 outbreak containment. In Veneto, where extensive testing was applied (3), the increase in COVID-19 mortality was milder than in the other regions, which initially clustered in a steeper relation between the number of tests and mortality. Accordingly, the proportion of positive tests was lower in Veneto than elsewhere, whereas the rate of daily increase in mortality in Emilia-Romagna, initially similar to that in Lombardy, declined when the rate of daily increase in the number of tests performed became steeper. Thus, whereas with its policy for extensive testing Veneto was efficaciously containing the spreading of the disease, Lombardy, Piedmont, and initially also Emilia-Romagna, were somehow chasing the virus, using tests more to confirm clinically plausible diagnoses than to contain the epidemics.

To estimate the spread of the disease, we used COVID-19 mortality instead of the number of positive tests, which depends heavily on the policy for testing: other factors being the same, the broader the criteria for testing, the wider the denominator, the lower the proportion of positive tests, and vice versa. A 7-day lag was allowed to identify deaths, as this is the minimal interval to attribute death to COVID-19 (2).

Being the first western country facing COVID-19 outbreak, Italy represents a living-lab to evaluate the effectiveness of practices to contrast it (8). In agreement with data from South Korea (9), a broader policy for swab testing, such as that applied in Veneto, appears to contribute successfully to contain COVID-19 threat.

## Data Availability

Data used in this study are in the public domain (https://github.com/pcm-dpc/COVID-19/tree/master/schede-riepilogative), being daily issued by the Departmet of Civil Protection of the Italian Government.

